# Effectiveness of Lifestyle Interventions for Glycemic Control among Adults with Type 2 Diabetes in West Africa: a Systematic Review and Meta-analysis

**DOI:** 10.64898/2026.05.16.26353078

**Authors:** Eugene Paa Kofi Bondzie, Nhyira Yaw Adjei-Banuah, Nana Efua Enyimayew Afun, Ellen Barnie Peprah, Yasmin Jahan, Tolib Mirzoev, Dina Balabanova, Irene Agyepong

## Abstract

Type 2 Diabetes (T2D) is a growing public health burden in West Africa, yet the effectiveness of lifestyle interventions for glycemic control in this region remains unclear. This systematic review and meta-analysis evaluated the impact of lifestyle interventions on Fasting Blood Glucose (FBG) and Glycated Hemoglobin (HbA1c) levels among adults with T2D in West Africa.

A systematic search of PubMed, Scopus, Africa Journals Online, and Cairn.info was conducted following PRISMA guidelines. Randomized controlled trials (RCTs) and quasi-experimental studies evaluating lifestyle interventions (physical activity, dietary modification, and combined/educational interventions) for glycemic control in adults with T2D in West Africa were included. Meta-analysis was performed via a random-effects model with restricted maximum likelihood (REML) estimation, using mean differences (MD) as the effect measure for both FBG and HbA1c outcomes. Heterogeneity was assessed via I² statistics, and sensitivity, subgroup, and meta-regression analyses were conducted to examine potential moderators of the observed heterogeneity.

Ten studies comprising 645 participants were included. Six studies reported FBG outcomes; however, two were excluded from the FBG meta-analysis due to missing control group post-test values and absence of a control group respectively, leaving four studies for pooling. A separate set of four studies contributed to the HbA1c meta-analysis. For FBG, lifestyle interventions were associated with reduction in FBG levels (pooled MD = −1.81 mmol/L, 95% CI: −2.33 to −1.30, p < 0.001), with moderate heterogeneity (I² = 50.76%). The certainty of evidence assessed using the GRADE approach was rated as low for FBG outcomes and very low for HbA1c outcomes, reflecting concerns about imprecision and inconsistency across studies.

Leave-one-out sensitivity analysis confirmed robustness of this finding, with estimates ranging from −1.707 to −2.087 mmol/L. Neither intervention duration nor sample size significantly moderated FBG effect sizes, with the model explaining approximately 15.7% of observed heterogeneity. For HbA1c, lifestyle interventions were also associated with reduction in HbA1c levels (pooled MD = −1.044%, 95% CI: −1.594 to −0.495, p = 0.0002), though heterogeneity was exceptionally high (I² = 98.08%), limiting interpretability of the pooled estimate. Exploratory meta-regression identified intervention duration and sample size as statistically associated with HbA1c effect size, though the model was saturated given the small number of studies and findings should not be interpreted as confirmatory evidence of moderation.

**Conclusion:** Lifestyle interventions, including supervised physical activity, dietary modification, and community-based diabetes education, were generally associated with improvements in glycemic control among adults with type 2 diabetes in West Africa. Evidence was more consistent for fasting blood glucose, while findings for HbA1c were highly heterogeneous and should be interpreted with caution. These results suggest potential benefit, but variability across studies highlights the need for more standardized and rigorously designed trials in the region.

## Introduction

Diabetes mellitus affects more than 537 million adults worldwide and remains a growing public health challenge, particularly in low- and middle-income countries (LMICs) (1). The World Health Organization reports that approximately 1.5 million deaths are directly attributable to diabetes annually, with developing regions facing significant challenges in timely diagnosis and effective management, which lead to high rates of microvascular and cardiovascular complications (2,3).

In West Africa, a sub-region comprising 16 developing economies, the burden of type 2 diabetes is compounded by rapid urbanization, shifting demographics, and constrained healthcare resources (4,5). The lingering effects of the COVID-19 pandemic have further strained health and other systems, exacerbating the difficulties in managing noncommunicable diseases (6,7). With the United Nations Sustainable Development Goal (SDG) 3.4 targeting a one-third reduction in premature noncommunicable disease mortality by 2030 (8), there is an urgent need to identify interventions that can bridge existing gaps in diabetes care.

Lifestyle interventions, particularly those focused on physical activity and medical nutrition therapy, have been identified as cornerstone strategies for glycemic control and improved cardiometabolic outcomes in patients with type 2 diabetes (9,10). Physical activity encompasses various activities, from daily movements such as walking and household chores to structured exercise programs such as aerobic and resistance training, with a minimum recommendation of 150 minutes of moderate-to-vigorous activity per week (10,11). Medical nutrition therapy, which is optimally delivered by registered dietitians, aims to regulate energy balance, optimize macronutrient distribution, and achieve individualized glycemic targets to prevent diabetes-related complications (12).

Although substantial evidence supports the effectiveness of these interventions in improving glycemic control in diverse populations (13–16), there is a paucity of research focusing specifically on West African populations. Prior reviews have included studies from low- and middle-income countries (LMICs), but the vast heterogeneity in cultural, socioeconomic, and health system contexts often limits the generalizability of their findings to West Africa (17). Furthermore, debates persist regarding the optimal design, intensity, and duration of lifestyle interventions, as well as the comparative effectiveness of individual-level versus community-level strategies (12,13,18).

Given these gaps, synthesizing the available evidence on the effectiveness of lifestyle interventions for glycemic control among adults with type 2 diabetes in West Africa is imperative. A rigorous synthesis will not only inform clinical practice and policy development in the region but also guide future research by highlighting areas that require further investigation.

## Objectives

1. To identify which individual-level/community-level lifestyle interventions (e.g., physical activity, nutrition therapy) are available for glycemic control among adults with type 2 diabetes in West Africa.
2. To explore the effectiveness of these lifestyle interventions in achieving glycemic control.

## Methods

Our systematic review and meta-analysis protocol was registered in the International Prospective Register for Systematic Reviews - PROSPERO, with registration number CRD42023435116 and published (19). We adhered to the PRISMA reporting guidelines throughout the review process(S1. File) (20).

Any modifications from the published protocol were made to enhance the review process; for example, we employed Covidence software for study screening (instead of Rayyan) owing to its enhanced collaborative features, and we used the Joanna Briggs Institute (JBI) critical appraisal tool (21) for nonrandomized studies rather than the ROBINS-I tool. These adjustments were implemented to improve efficiency and rigor.

### Criteria for Considering Studies

We used the population, intervention, comparison, outcome, and study design (PICOS) framework to determine study inclusion.

### Population

Studies were included if they involved adults (≥18 years) residing in West Africa with a diagnosis of type 2 diabetes. Studies focusing on type 1 diabetes, pediatric, or gestational diabetes were excluded.

### Intervention

We considered all lifestyle interventions related to physical activity and nutrition. The physical activity interventions included low-, moderate-, and high-intensity exercises, including structured exercise regimens as well as non sedentary everyday movements (e.g., walking, gardening, housework), provided that they were delivered within a defined regimen and appropriately measured (10). Nutrition interventions include various dietary modifications, such as vegetarian, low-carbohydrate, low-fat, or plant-based diets (12). Interventions were classified as either individual-level (e.g., one-on-one counseling, structured education programs) or community-level (e.g., public awareness campaigns, community-based physical activity programs).

### Control

The control condition was defined as usual care or no intervention. Usual care was defined as the standard diabetes care provided in each study, which typically included routine medical consultations, pharmacological treatment, and general lifestyle advice. However, there was some variation across studies in the specifics of usual care. In all cases, the control groups did not receive structured lifestyle interventions such as supervised physical activity, dietary counseling, or community-based diabetes education.

### Outcome

The primary outcome was glycemic control, which was measured by changes in Glycated Hemoglobin (HbA1c) levels and Fasting Blood Glucose (FBG) levels. Although some researchers have raised concerns about the cost and biological variability of HbA1c measurements (22), it remains a widely accepted metric for assessing glycemic improvement in clinical trials (23). An intervention was deemed effective if it resulted in a clinically significant reduction in HbA1c of at least 5 mmol/mol (or 0.5%); reductions below this threshold, no change, or an increase were considered noneffective (24,25). A clinically significant reduction in FBG is generally considered to be in the range of approximately 20–30 mg/dL (1.1–1.7 mmol/L) from baseline values (10).

### Studies

Eligible study designs included Randomized Controlled Trials (RCTs) and quasi experimental studies (e.g., pretest-posttest designs, nonequivalent control group designs, and controlled observational studies) that aimed to establish causal relationships between the intervention and glycemic outcomes.

### Search strategy

We searched four electronic databases including PubMed, Scopus, Africa Journals Online, and Cairn.info for articles published between 2000 and March 2026. Additionally, relevant websites of government agencies and nongovernmental organizations (e.g., PATH, Sante Diabete) were searched for program reports, evaluations, and pertinent publications, and clinical trial registries were examined for ongoing or recently completed trials. To ensure comprehensiveness, we also screened the reference lists and bibliographies of all included studies.

Our search strategy combined keywords and medical subject headings (MeSH) related to “diabetes”, “lifestyle modification”, “physical activity”, “nutrition” and their synonyms. The full search strategy is provided in S2.File. Searches were restricted to studies published in English or French, as these are the most widely used for scholarly publications and reports within the West Africa sub-region. A search alert was established to capture new studies during the review process.

### Study Selection and Management

Two reviewers independently screened titles and abstracts via the Covidence software platform to manage the search results and remove duplicates. Discrepancies were resolved by a third reviewer. The full-text articles were then independently assessed against the inclusion criteria, with any disagreements resolved through consensus. The study selection process is detailed in S3.File.

### Risk-of-bias assessment

The risk of bias of selected RCTs was evaluated via the Cochrane Collaboration tool (26). Two independent reviewers conducted the assessments, categorizing studies as low risk, high risk, or unclear based on six domains: selection bias (random sequence generation, allocation concealment); performance bias (blinding of participants and personnel); detection bias (blinding of outcome assessment); attrition (incomplete outcome data); reporting bias (incomplete reporting); and other bias. For nonrandomized studies, we employed the JBI critical appraisal tool. A third reviewer resolved any discrepancies.

### Data Extraction and Management

Data were independently extracted by two reviewers via a predesigned Microsoft Excel form. The following data were collected: first author’s last name, publication year, country, study setting, participant characteristics (sample size and mean age), details of the intervention (type, frequency, and duration), characteristics of the control group, pre- and postintervention FBG/HbA1c levels, additional outcomes, and the authors’ conclusions. We contacted the corresponding authors of relevant studies for which data related to study methods and outcomes were unclear or missing via emails, providing a 4-week timeline for response. In cases where authors did not respond, we proceeded with data synthesis and clearly reported the missing information and its potential impact on the overall findings in the limitations section. The absence of this information does not compromise the core dataset necessary to reproduce our findings.

### Data Synthesis

The effect of interventions on FBG and HbA1c was estimated using mean differences (MD) with 95% confidence intervals, expressed in mmol/L and percentage points respectively, to facilitate clinical interpretability. Mean differences were calculated as intervention group minus control group, such that negative values indicate greater reductions in the intervention group relative to control. Where necessary, effect estimates were converted to a common metric. Measures of precision were reported as 95% confidence intervals, calculated on the basis of the number of participants per treatment group (27). When both pre- and post-intervention measures were available, endpoint data and their standard deviations were used.

Heterogeneity among studies was assessed via the I² statistic. Owing to anticipated heterogeneity, a random-effects model with restricted maximum likelihood (REML) estimation was applied for all meta-analyses. Forest plots were generated to visualize the pooled effect sizes and the extent of heterogeneity. Sensitivity analyses were conducted using a leave-one-out approach to explore the influence of individual studies on the overall pooled estimate. Subgroup analyses were performed stratified by intervention type and duration. Meta-regression was performed to examine whether intervention duration and sample size explained between-study variability in effect sizes. All meta-analyses were performed using STATA (version 18).

### Assessment of Certainty of Evidence

The certainty of the body of evidence for each outcome was assessed using the Grading of Recommendations, Assessment, Development and Evaluations (GRADE) approach (28). For each outcome, evidence was assessed across five domains: risk of bias, inconsistency, indirectness, imprecision, and publication bias. Evidence from randomised controlled trials begins at high certainty and is downgraded based on the presence and severity of concerns within each domain. A Summary of Findings table was generated to present the certainty ratings alongside pooled effect estimates for each outcome.

## Results

A total of 2,893 records were identified through searches of databases and trial registers. After 538 duplicates were removed, 2,355 unique records were screened by title and abstract by two independent reviewers, resulting in the exclusion of 2,331 records. Twenty-four full-text articles were then assessed, and 14 studies were excluded because of unsuitable settings, interventions, study designs, or patient populations. Ultimately, 10 studies involving 645 participants were included in the systematic review and meta-analysis (Fig 1).

**Fig 1.**
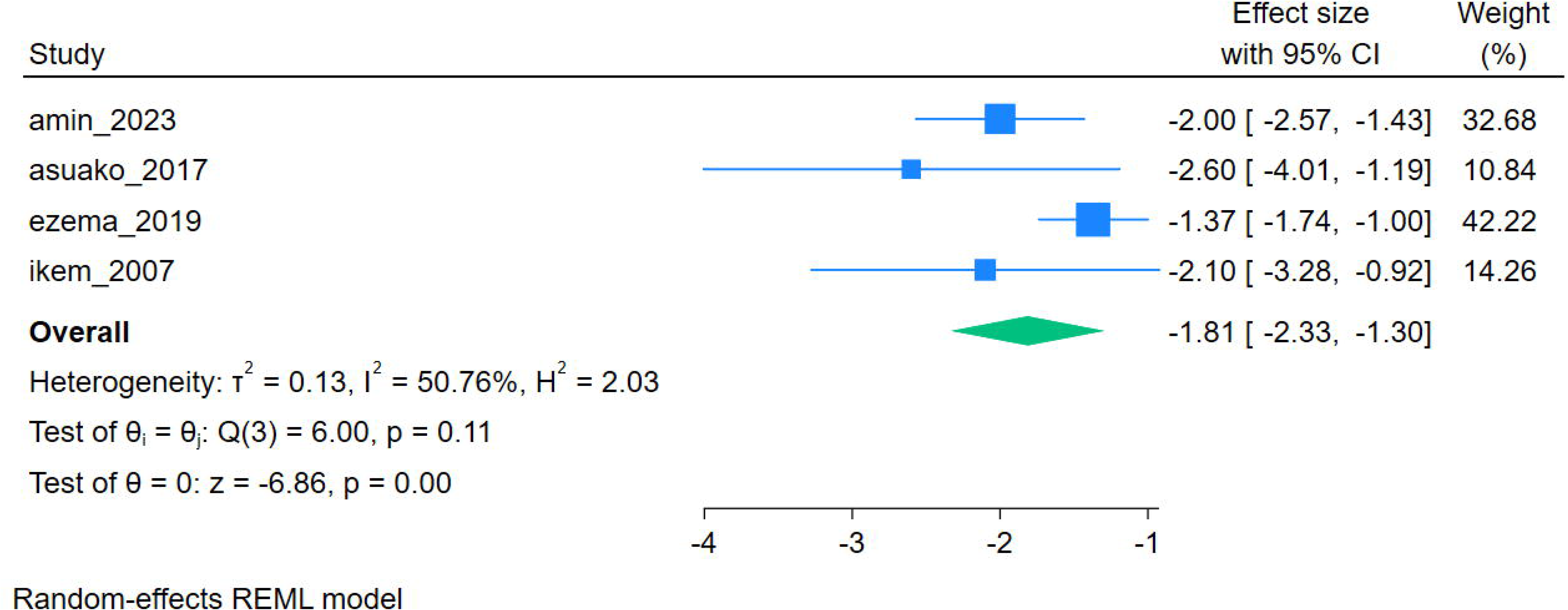
PRISMA diagram with the flowchart of study screening and inclusion.

Most studies were from Nigeria (n = 7), followed by Ghana (n = 2) and Mali (n = 1). Nine studies were RCTs (29–37), and one was a experimental pretest–posttest study without a control group (38). Interventions were categorized as follows: six studies focused on physical activity interventions (29–33, 38), one evaluated a conventional dietary intervention (37), one evaluated a herbal supplement intervention (34), and two assessed combined or educational interventions that integrated elements of both physical activity and dietary modifications (35, 36). The herbal supplement study was retained for completeness in the systematic review but was not considered part of the core lifestyle intervention category and was not included in pooled quantitative interpretations, given its distinct mechanism of action. Nine studies reported significant effects on glycemic control, whereas one did not (34).

The risk of bias was evaluated via the Cochrane Collaboration tool for RCTs and the JBI critical appraisal tool for quasi experimental studies. Overall, eight studies were rated as having a low risk of bias (29,31,33–37), one was judged as unclear (32) and one study was classified as high risk (38) due to incomplete follow-up and the absence of a control group. A summary of the study characteristics and risk-of-bias assessments is provided in Table 1. A complete table of study characteristics can be found in S1 Table.

**Table 1.**
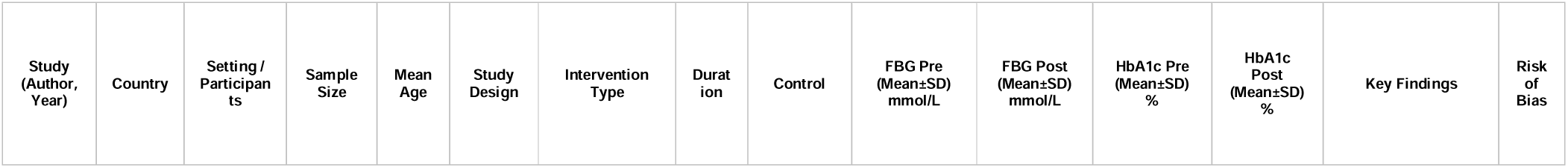
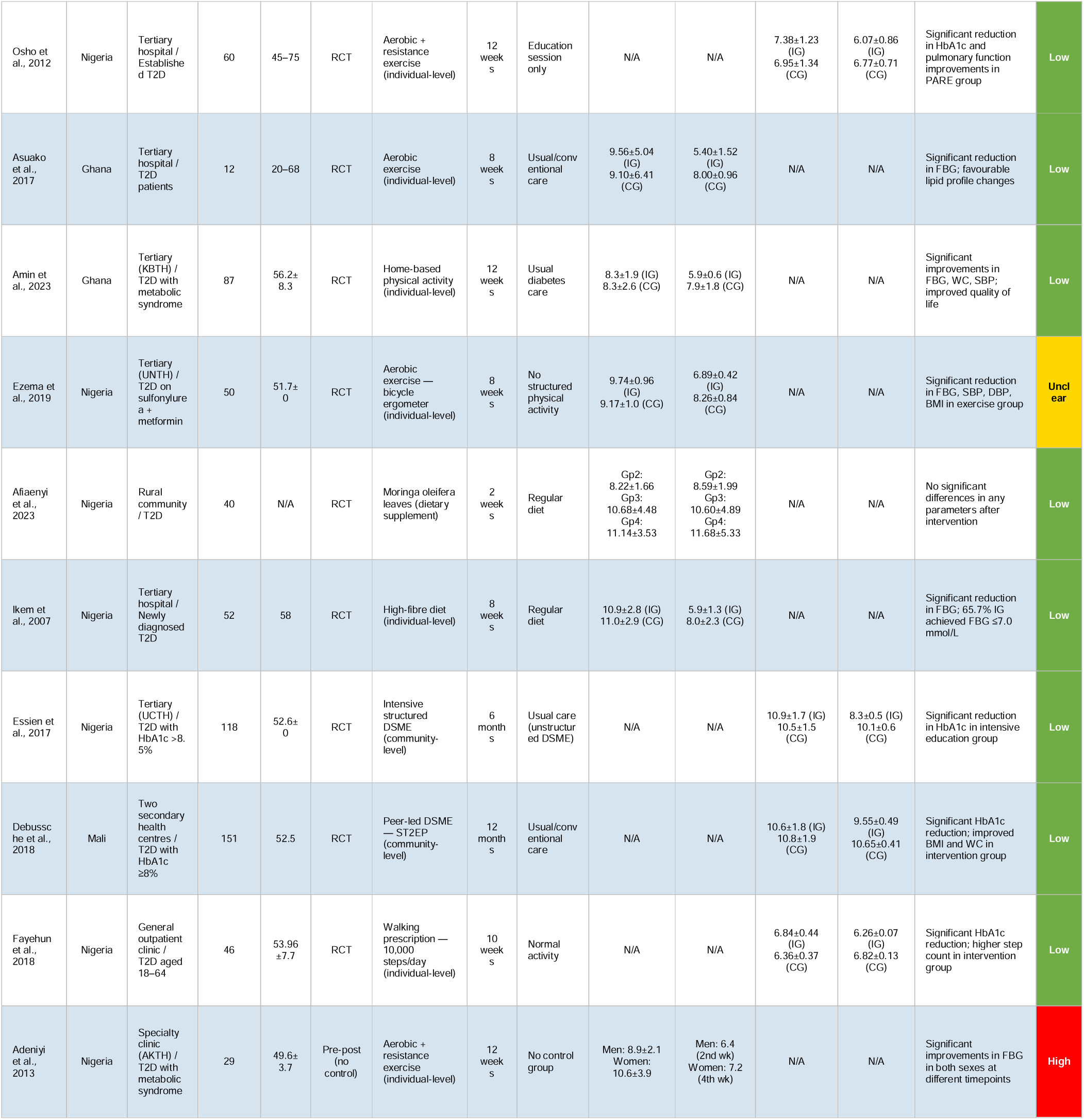
Summary of the study characteristics and risk-of-bias assessments.

| Study (Author, Year) | Country | Setting / Participants | Sample Size | Mean Age | Study Design | Intervention Type | Duration | Control | FBG Pre (Mean±SD) mmol/L | FBG Post (Mean±SD) mmol/L | HbA1c Pre (Mean±SD) % | HbA1c Post (Mean±SD) % | Key Findings | Risk of Bias |
| --- | --- | --- | --- | --- | --- | --- | --- | --- | --- | --- | --- | --- | --- | --- |
| Osho et al., 2012 | Nigeria | Tertiary hospital / Established T2D | 60 | 45–75 | RCT | Aerobic + resistance exercise (individual-level) | 12 weeks | Education session only | N/A | N/A | 7.38±1.23 (IG)<br>6.95±1.34 (CG) | 6.07±0.86 (IG)<br>6.77±0.71 (CG) | Significant reduction in HbA1c and pulmonary function improvements in PARE group | Low |
| Asuako et al., 2017 | Ghana | Tertiary hospital / T2D patients | 12 | 20–68 | RCT | Aerobic exercise (individual-level) | 8 weeks | Usual/conventional care | 9.56±5.04 (IG)<br>9.10±6.41 (CG) | 5.40±1.52 (IG)<br>8.00±0.96 (CG) | N/A | N/A | Significant reduction in FBG; favourable lipid profile changes | Low |
| Amin et al., 2023 | Ghana | Tertiary (KBTB) / T2D with metabolic syndrome | 87 | 56.2±8.3 | RCT | Home-based physical activity (individual-level) | 12 weeks | Usual diabetes care | 8.3±1.9 (IG)<br>8.3±2.6 (CG) | 5.9±0.6 (IG)<br>7.9±1.8 (CG) | N/A | N/A | Significant improvements in FBG, WC, SBP, improved quality of life | Low |
| Ezema et al., 2019 | Nigeria | Tertiary (UNTH) / T2D on sulfonylurea + metformin | 50 | 51.7±0 | RCT | Aerobic exercise — bicycle ergometer (individual-level) | 8 weeks | No structured physical activity | 9.74±0.96 (IG)<br>9.17±1.0 (CG) | 6.89±0.42 (IG)<br>8.26±0.84 (CG) | N/A | N/A | Significant reduction in FBG, SBP, DBP, BMI in exercise group | Unclear |
| Afiyaeni et al., 2023 | Nigeria | Rural community / T2D | 40 | N/A | RCT | Moringa oleifera leaves (dietary supplement) | 2 weeks | Regular diet | Gp2: 8.22±1.66<br>Gp3: 10.68±4.48<br>Gp4: 11.14±3.53 | Gp2: 8.59±1.99<br>Gp3: 10.60±4.89<br>Gp4: 11.68±5.33 | N/A | N/A | No significant differences in any parameters after intervention | Low |
| Ikem et al., 2007 | Nigeria | Tertiary hospital / Newly diagnosed T2D | 52 | 58 | RCT | High-fibre diet (individual-level) | 8 weeks | Regular diet | 10.9±2.8 (IG)<br>11.0±2.9 (CG) | 5.9±1.3 (IG)<br>8.0±2.3 (CG) | N/A | N/A | Significant reduction in FBG; 65.7% IG achieved FBG ≤7.0 mmol/L | Low |
| Essien et al., 2017 | Nigeria | Tertiary (UCTH) / T2D with HbA1c >8.5% | 118 | 52.6±0 | RCT | Intensive structured DSME (community-level) | 6 months | Usual care (unstructured DSME) | N/A | N/A | 10.9±1.7 (IG)<br>10.5±1.5 (CG) | 8.3±0.5 (IG)<br>10.1±0.6 (CG) | Significant reduction in HbA1c in intensive education group | Low |
| Debussche et al., 2018 | Mali | Two secondary health centres / T2D with HbA1c ≥8% | 151 | 52.5 | RCT | Peer-led DSME — ST2EP (community-level) | 12 months | Usual/conventional care | N/A | N/A | 10.6±1.8 (IG)<br>10.8±1.9 (CG) | 9.55±0.49 (IG)<br>10.65±0.41 (CG) | Significant HbA1c reduction; improved BMI and WC in intervention group | Low |
| Fayehun et al., 2018 | Nigeria | General outpatient clinic / T2D aged 18–64 | 46 | 53.96±7.7 | RCT | Walking prescription — 10,000 steps/day (individual-level) | 10 weeks | Normal activity | N/A | N/A | 6.84±0.44 (IG)<br>6.36±0.37 (CG) | 6.26±0.07 (IG)<br>6.82±0.13 (CG) | Significant HbA1c reduction; higher step count in intervention group | Low |
| Adeniyi et al., 2013 | Nigeria | Specialty clinic (AKTH) / T2D with metabolic syndrome | 29 | 49.6±3.7 | Pre-post (no control) | Aerobic + resistance exercise (individual-level) | 12 weeks | No control group | Men: 8.9±2.1<br>Women: 10.6±3.9 | Men: 6.4 (2nd wk)<br>Women: 7.2 (4th wk) | N/A | N/A | Significant improvements in FBG in both sexes at different timepoints | High |

### Fasting Blood Glucose (FBG) Outcomes

A total of six studies reported on fasting blood glucose (FBG) outcomes. However, two were excluded from the meta-analysis: Afiaenyi et al. (34) was excluded due to missing control group post-test FBG values, which precluded the calculation of a valid mean difference and Adeniyi et al. (38) was excluded due to the absence of a control group. The remaining four studies (R.T. Ikem et al., Asuako et al., Amin et al., and Ezema et al.) (37, 30, 31, 32) were included in the meta-analysis. All included studies evaluated the effect of lifestyle-based interventions, including dietary modification and/or physical activity, compared with usual care or control conditions. Across studies, reductions in FBG were consistently observed in the intervention groups relative to controls, with individual study mean differences ranging from −1.37 to −2.60 mmol/L.

The meta-analysis using a random-effects model with restricted maximum likelihood (REML) estimation suggests that lifestyle interventions are associated with reductions in fasting blood glucose levels compared with control conditions. The pooled mean difference was −1.81 mmol/L (95% CI: −2.33 to −1.30, p < 0.001), indicating a statistically significant improvement (Fig 2). This reduction exceeds the predefined threshold of 1.1 mmol/L considered clinically meaningful for FBG improvement.

**Fig 2.**
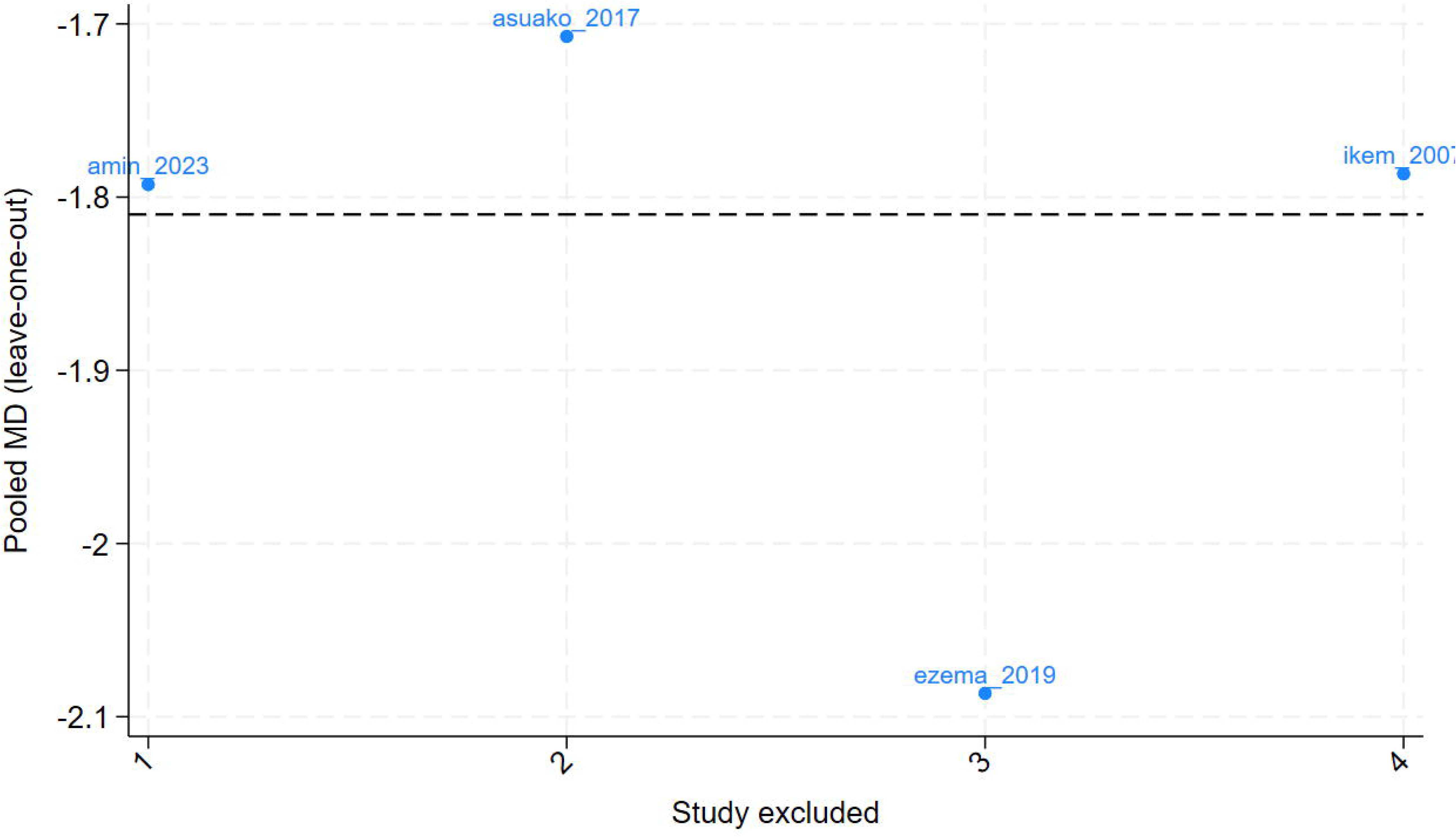
Forest Plot of effect of interventions of FBG.

Although all studies demonstrated effects in the same direction, moderate heterogeneity was observed across studies (I² = 50.76%, Cochran’s Q = 6.00, p = 0.1117; τ² = 0.1301), suggesting that variability in effect estimates may partly reflect real differences in study populations, intervention delivery, or contextual factors rather than chance alone.

Visual inspection of the funnel plot did not show clear evidence of asymmetry, although interpretation remains limited due to the small number of included studies.

Leave-one-out sensitivity analysis showed that the pooled effect remained stable across all iterations. When R.T. Ikem et al. was excluded, the pooled effect was −1.787 mmol/L. Excluding Asuako et al. produced a pooled estimate of −1.707 mmol/L. Removal of Amin et al. resulted in a pooled estimate of −1.793 mmol/L, while exclusion of Ezema et al. yielded a pooled estimate of −2.087 mmol/L (Fig 3). Across all iterations, the direction of effect remained consistently negative and statistically significant, with only minor changes in magnitude and heterogeneity.

**Fig 3.**
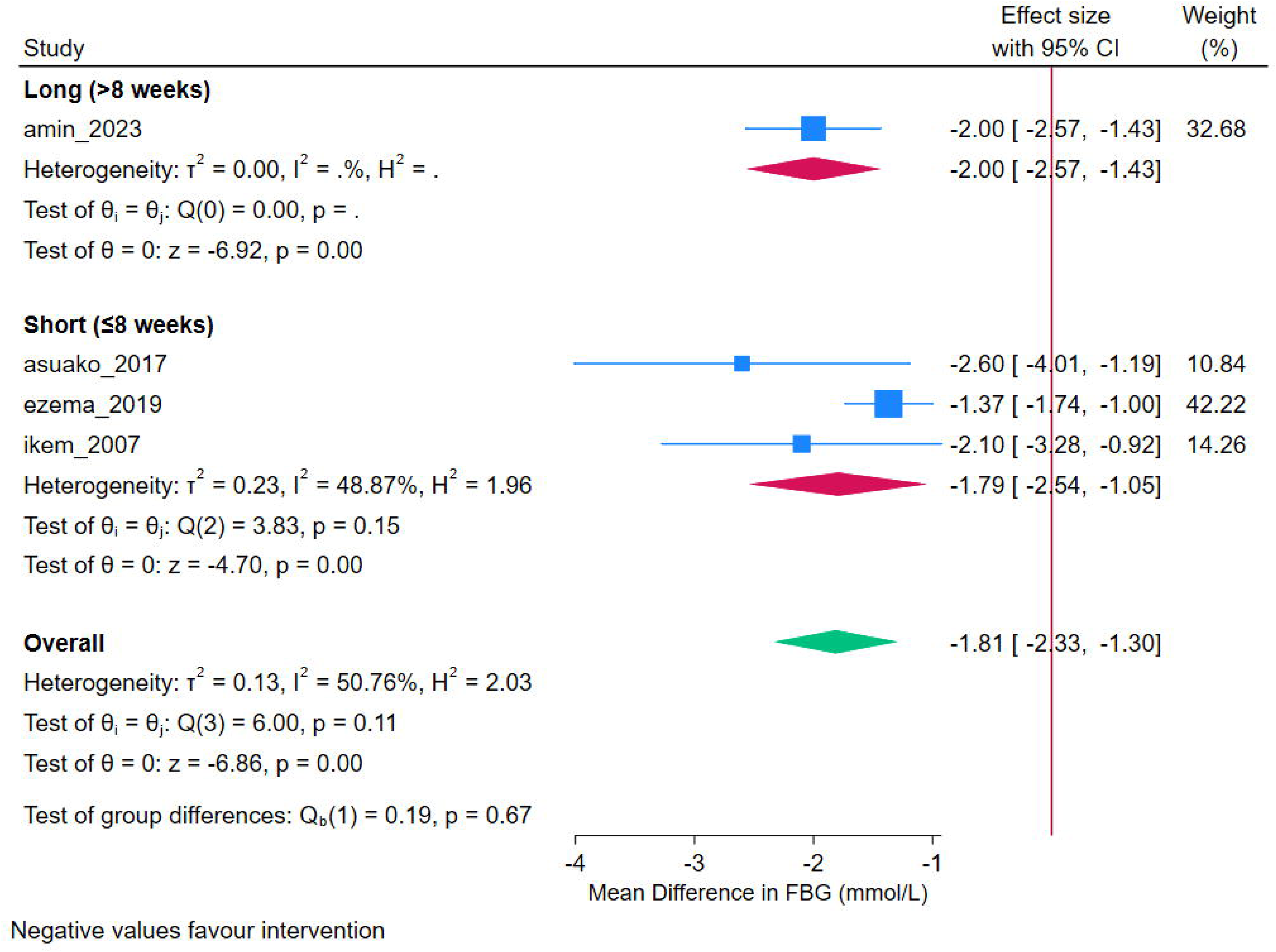
Leave-one-out sensitivity analysis of FBG effect sizes.

Overall, no single study exerted a disproportionate influence on the pooled effect, and the association between lifestyle interventions and improved FBG appears robust.

Subgroup analysis stratified by intervention type revealed that all intervention types demonstrated reductions in FBG compared with control conditions. Two studies evaluated aerobic exercise interventions, yielding a pooled mean difference of −1.79 mmol/L (95% CI: −2.93 to −0.65), with moderate within-subgroup heterogeneity (I² = 63.7%). One study assessed a high-fibre dietary intervention, reporting a mean difference of −2.10 mmol/L (95% CI: −3.28 to −0.93), and one study evaluated a home-based physical activity programme, reporting a mean difference of −2.00 mmol/L (95% CI: −2.57 to −1.43). The test for subgroup differences was not statistically significant (Qb = 0.15, p = 0.927), indicating no evidence of differential effects by intervention type (Fig 4). Given the small number of included studies and the uneven distribution across subgroups, these analyses were strictly exploratory in nature and no inferential conclusions should be drawn regarding effect modification.

**Fig 4.**
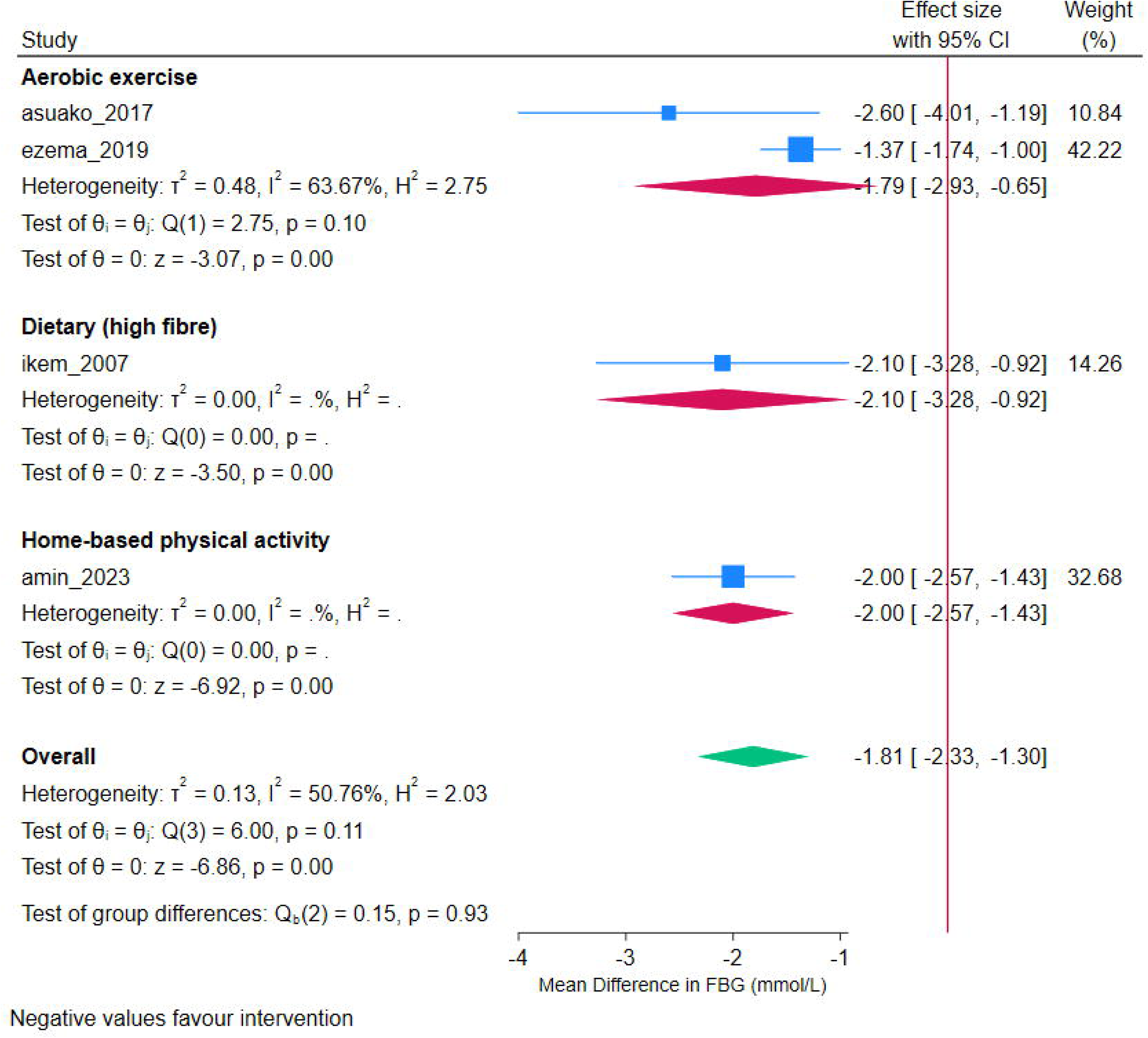
Subgroup analysis of FBG effect sizes stratified by intervention duration.

Subgroup analysis stratified by intervention duration revealed that short-duration interventions (≤8 weeks; three studies) yielded a pooled mean difference of −1.79 mmol/L (95% CI: −2.54 to −1.05), indicating a statistically significant reduction in FBG. The single long-duration study (>8 weeks) reported a mean difference of −2.00 mmol/L (95% CI: −2.57 to −1.43). The test for subgroup differences was not statistically significant (Qb = 0.19, p = 0.665) (Fig 5). Given the imbalance in subgroup composition and the limited number of studies, conclusions regarding effect modification by intervention duration are exploratory.

**Fig 5.**
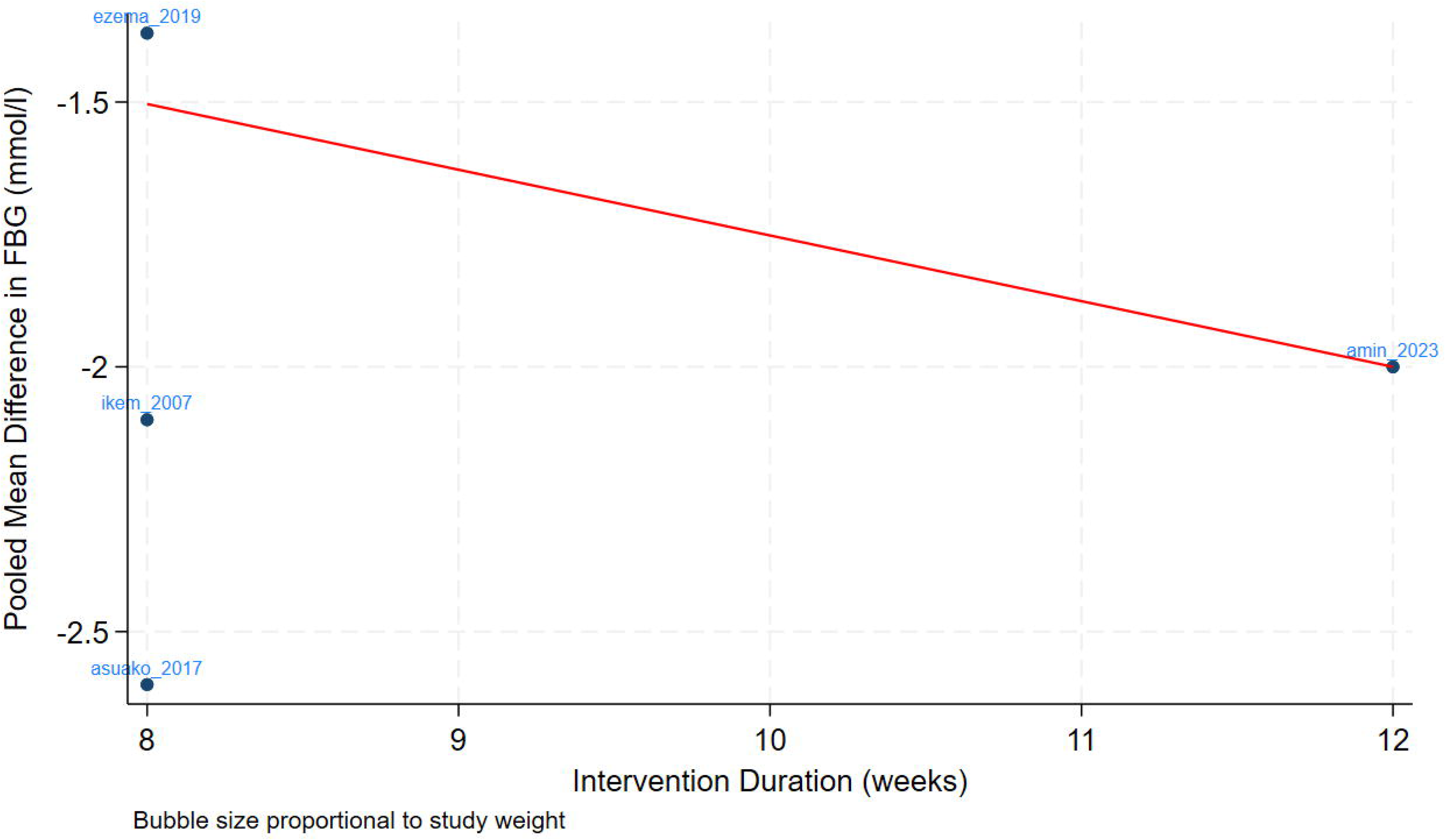
Subgroup analysis of FBG effect sizes stratified by intervention type.

Meta-regression was performed to examine whether intervention duration and sample size explained between-study variability in FBG effect sizes (Fig 6). Intervention duration showed a negative but non-significant association with effect size (β = −0.35, p = 0.194), suggesting a tendency toward greater reductions in FBG with longer interventions, although this relationship did not reach statistical significance. Sample size was not significantly associated with effect size (β = 0.026, p = 0.238). The model accounted for approximately 15.7% of the observed heterogeneity, indicating limited explanatory power of the included covariates. Residual heterogeneity remained, suggesting that unmeasured study-level factors such as intervention intensity, adherence, and contextual determinants likely contribute to the variability observed across studies.

**Fig 6.**
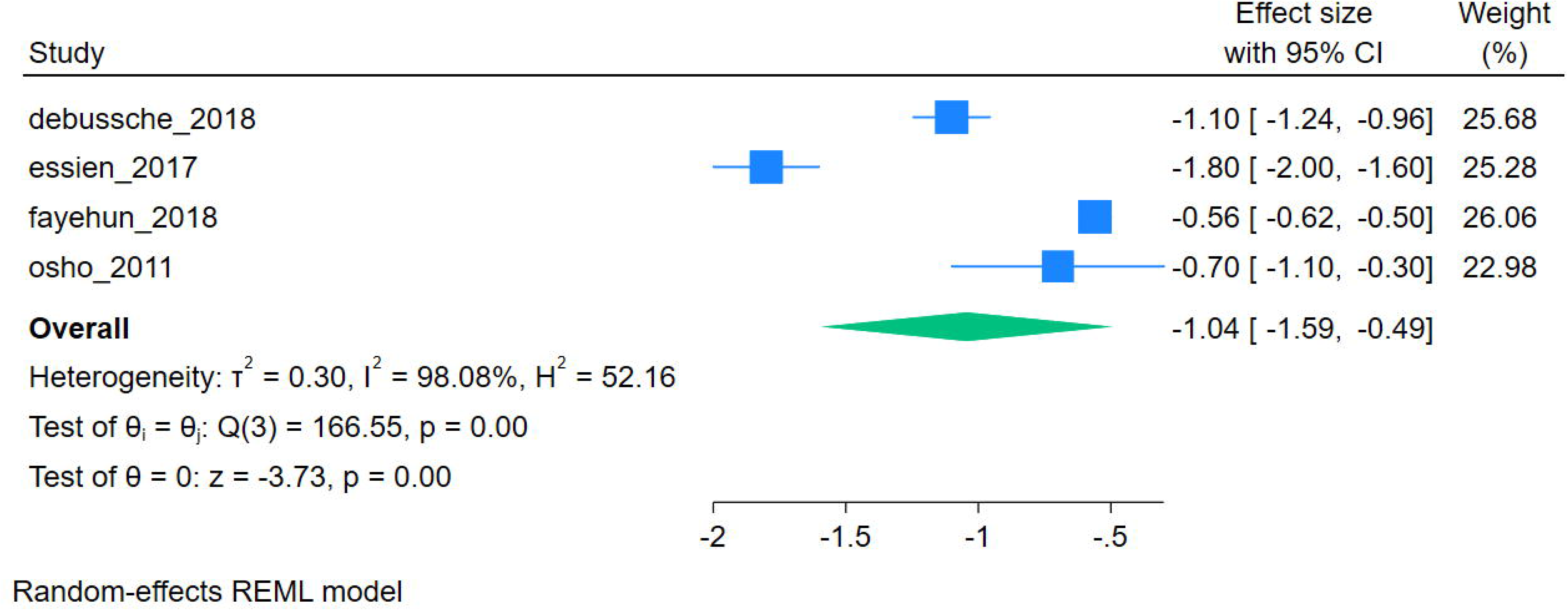
Scatter plot showing the association between intervention duration, sample size, and pooled mean difference in FBG.

### Glycated Hemoglobin (HbA1c) Outcomes

A separate meta-analysis of four studies reporting on HbA1c (Osho et al., Essien et al., Debussche et al., and Fayehun et al.) (29, 35, 36, 33) yielded a pooled mean difference of −1.044% (95% CI: −1.594 to −0.495, p = 0.0002), favouring lifestyle interventions over usual care (Fig 7). However, heterogeneity was exceptionally high (I² = 98.08%, Cochran’s Q = 166.55, p < 0.001; τ² = 0.3005), indicating that nearly all variability in effect estimates reflects genuine differences among studies rather than sampling error. At this level of inconsistency, the pooled mean difference should be interpreted as a descriptive average rather than a precise estimate of a common intervention effect. The observed variability suggests that intervention effects are highly context-specific, and conclusions regarding the magnitude of effect should be made with caution. Leave-one-out sensitivity analysis showed that the pooled effect changed modestly across iterations, with estimates ranging from −0.791 to −1.216% (Fig 8). The overall pooled estimate was −1.044%. When Debussche et al. was excluded, the pooled effect was −1.024%. Excluding Essien et al. produced the largest shift, yielding a pooled estimate of −0.791%, reflecting the relatively strong influence of this study on the overall effect. Removal of Fayehun et al. resulted in a pooled estimate of −1.216%, while exclusion of Osho et al. yielded a pooled estimate of −1.148%. Across all iterations, the direction of effect remained consistently negative and statistically significant, although the exclusion of Essien et al. produced a notable reduction in the magnitude of the pooled effect, suggesting that this study exerts relatively greater influence on the overall estimate compared with the other included studies.

**Fig 7.**
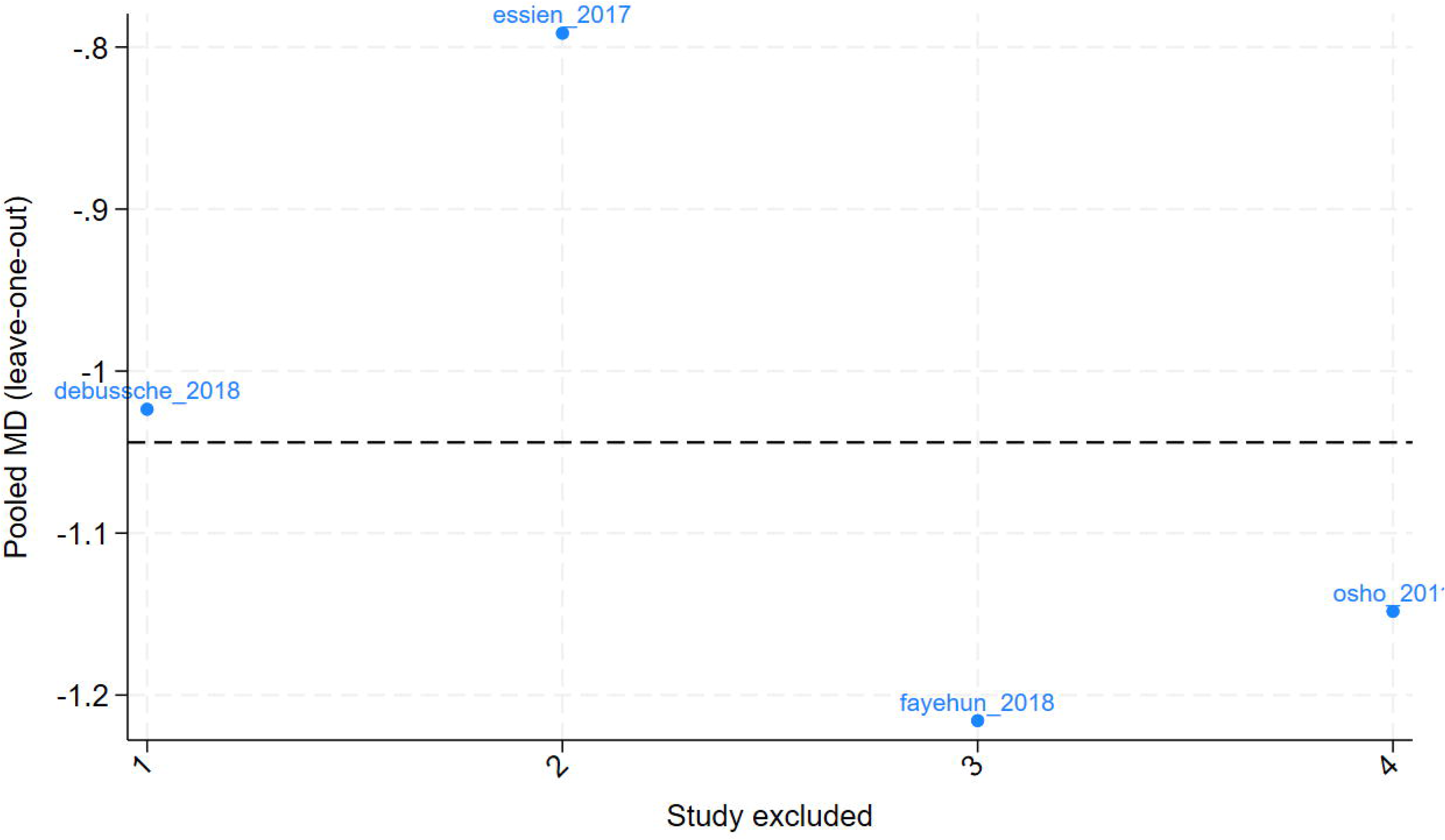
Forest Plot of the effect of interventions on HbA1c.

**Fig 8.**
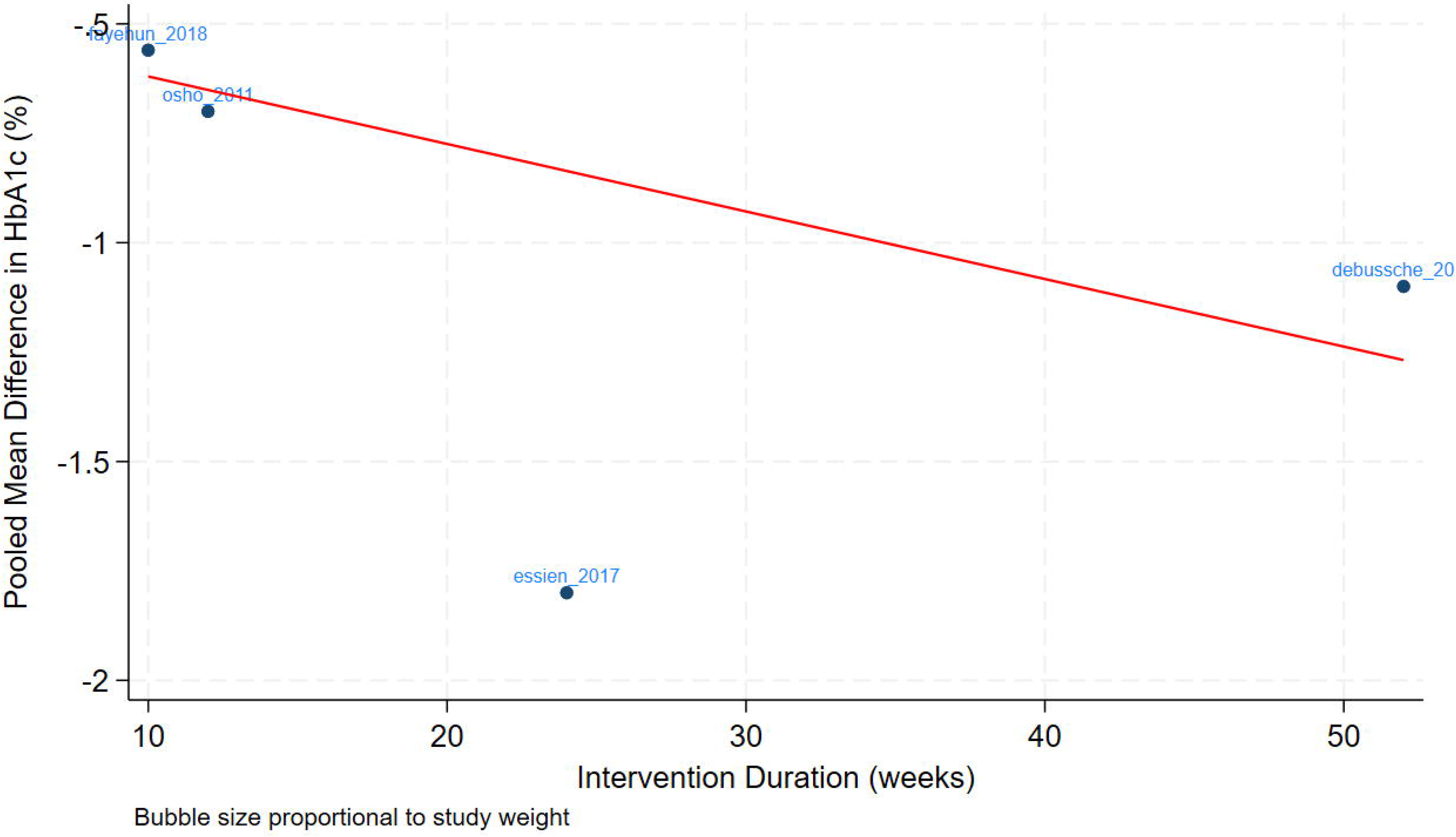
Leave-one-out sensitivity analysis of HbA1c effect sizes.

Subgroup analysis of HbA1c outcomes stratified by sample size was not performed, as the small number of included studies precluded a meaningful between-group comparison. With only four studies available, dividing the sample into subgroups would result in subgroups of two studies each, providing insufficient data to draw reliable conclusions. This reflects a limitation of the current evidence base, and future reviews incorporating a larger number of studies will be better positioned to examine the moderating role of sample size on HbA1c outcomes. Similarly, subgroup analysis stratified by intervention type could not be meaningfully performed for HbA1c outcomes. Although the four included studies could be broadly categorised into education-based and physical activity interventions, with only two studies per subgroup, pooled estimates would be statistically unreliable and the between-group comparison would lack sufficient power to detect meaningful differences. This analysis will be more informative in future reviews with a larger evidence base.

Meta-regression examining intervention duration and sample size as potential moderators was conducted as an exploratory analysis (Fig 9). The coefficient for intervention duration was 0.058 (95% CI: 0.043 to 0.073, p < 0.001) and for sample size was −0.028 (95% CI: −0.034 to −0.023, p < 0.001). With only four studies and two covariates, however, the model is saturated, meaning there are no remaining degrees of freedom for residual heterogeneity. The R² of 100% therefore reflects a mathematical property of the saturated model rather than empirical evidence that these two variables fully explain the observed heterogeneity. A larger body of studies would be required to draw meaningful conclusions about the role of intervention duration and sample size in explaining variability in HbA1c outcomes.

**Fig 9.**
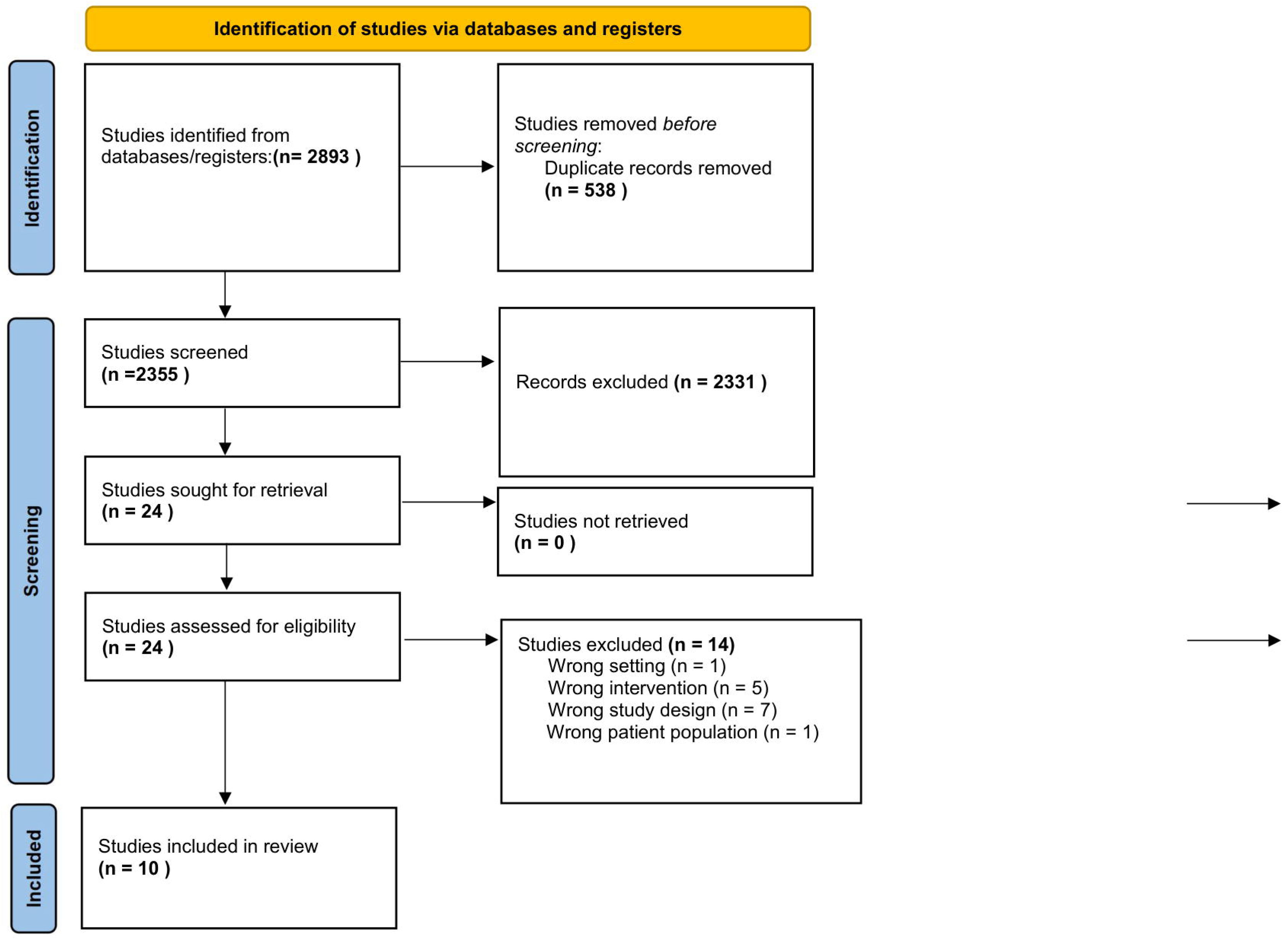
Scatter plot showing the association between intervention duration, sample size, and pooled mean difference in HbA1c.

The certainty of evidence was assessed using the GRADE approach for both primary outcomes (Table 2). For FBG outcomes, the evidence was rated as low certainty. Although all four included studies were RCTs with low risk of bias, the evidence was downgraded by one level for inconsistency due to moderate heterogeneity (I² = 50.76%) and by one level for imprecision given the small number of participants and wide confidence intervals. For HbA1c outcomes, the evidence was rated as very low certainty. The evidence was downgraded by two levels for inconsistency due to exceptionally high heterogeneity (I² = 98.08%) and by one level for imprecision. These ratings indicate that while the observed effects are statistically significant, considerable uncertainty remains regarding the true magnitude of benefit, and future well-designed trials are needed to strengthen the evidence base.

**Table 2.** Certainty of evidence for outcomes measures using GRADE.

| Outcome | No. Studies (Participants) | Design | Risk of Bias | Inconsistency | Indirectness | Imprecision | Publication Bias | GRADE Certainty |
| --- | --- | --- | --- | --- | --- | --- | --- | --- |
| Fasting Blood Glucose (FBG) [mmol/L] | 4 RCTs (n=209) | RCTs | Not serious (all 4 low ROB) | Serious (I<sup>2</sup>=50.76%) | Not serious (West Africa) | Serious (wide CI; n<400) | Undetected (n=4) | LOW |
| Glycated Haemoglobin (HbA1c) [%] | 4 RCTs (n=375) | RCTs | Not serious (all 4 low ROB) | Very serious (I<sup>2</sup>=98.08%) | Not serious (West Africa) | Serious (wide CI; n<400) | Undetected (n=4) | VERY LOW |

GRADE certainty: High (green), Moderate (amber), Low (orange), Very Low (dark red) | Domains: Not serious (green), Serious (yellow), Very serious (red)

## Discussion

Our systematic review and meta[analysis indicate that structured lifestyle interventions such as supervised physical activity, dietary counseling, or community-based diabetes education are associated with improvements in glycemic control among adults with type 2 diabetes in West Africa, although effects varied substantially across studies. The pooled mean difference for FBG (MD = −1.81 mmol/L) and HbA1c (MD = −1.044%) suggest potential clinically meaningful improvements, when these interventions are added to usual care, which typically includes routine medical consultations, pharmacological treatment, and general lifestyle advice. These findings align with evidence from other regions and sub-regions.

For example, regarding supervised physical activity interventions, our review of West African studies revealed positive effects on glycemic control. Osho et al (29) implemented a 12-week aerobic and resistance exercise program in Nigeria, resulting in significant reductions in FBG and improvements in pulmonary function. Similarly, Asuako et al (30) reported that an 8-week aerobic exercise regimen in Ghana led to marked decreases in FBG and favorable lipid profile changes. These outcomes are consistent with evidence from other LMICs; for instance, Sagastume et al (17) found in a systematic review that structured exercise training significantly reduces HbA1c levels in individuals with type 2 diabetes. Similarly, a review by Colberg et al (10) on studies from the United States and other high-income countries supported the role of physical activity in improving glycemic control and insulin sensitivity.

In relation to dietary interventions, our findings from R.T Ikem et al (37), a controlled trial in Nigeria comparing a high-fiber diet to a conventional diet over eight weeks leading to significant improvements in FBG and lipid profiles among participants adhering to the high-fiber regimen, aligns with studies from other LMICs. Meta-analysis from O’Donoghue et al (39), found that dietary modifications, including plant-based diets, high fibre diet and carbohydrate control, significantly reduced HbA1c levels in adults with type 2 diabetes in low- and middle-income settings.

Additionally, pooled studies (40) from higher income settings among European and North American populations suggest that dietary patterns emphasizing plant-based foods, high fibre-diet and healthy fats contribute to better glycemic control and cardiovascular risk reduction.

Our analysis of community-level interventions also supports global findings. Essien et al (35) evaluated a six-month intensive diabetes self-management education program in Nigeria and found that, compared with conventional approaches, structured education led to significant reductions in HbA1c. Likewise, Debussche et al. (36) reported that a 12-month peer-led diabetes self-management programme in Mali resulted in improved glycemic and anthropometric outcomes. These findings are in line with systematic reviews conducted in the United Kingdom and the United States, where Attridge et al (41), concluded that health education significantly enhances glycemic control in ethnic minority groups with type 2 diabetes. Similarly, O’Donoghue et al (39) found that community-based and culturally appropriate diabetes education programs in LMICs significantly enhanced glycemic control and self-management behaviors.

The pooled effect sizes in our meta-analysis indicate that lifestyle targeted interventions (supervised physical activity, diet and community-based health education) show beneficial effects in reducing both FBG and HbA1c levels among West African populations. However, substantial variability was observed across studies, meaning that the effectiveness of these interventions differed significantly between studies, with some reporting stronger effects while others showed weaker or nonsignificant effects. This variability appears to stem from real differences in study characteristics rather than random variation. Our subgroup analyses revealed that studies with larger sample sizes yielded more consistent and statistically significant effect sizes with minimal variability than did smaller studies. This observation is consistent with the notion that larger studies provide more robust estimates of intervention effects. However, further analysis revealed that sample size alone did not explain the variability in the observed effects.

Additionally, meta-regression findings differed between outcomes. For FBG outcomes, neither intervention duration nor sample size was identified as a significant moderator of effect size, suggesting that contextual factors such as cultural beliefs, healthcare access, and socioeconomic conditions may have influenced effectiveness more than structural intervention characteristics. For HbA1c outcomes, exploratory meta-regression suggested that longer intervention duration was associated with relatively smaller reductions in HbA1c, while larger studies tended to show greater reductions. One possible explanation for this pattern is that the HbA1c studies in this review were predominantly longer-term, structured educational and community-based interventions, whereas the FBG studies were largely shorter-term exercise programmes, and these differences in intervention design and study context may partly account for the divergent meta-regression findings across outcomes. From a biological standpoint, HbA1c reflects average glycemic control over two to three months, meaning that shorter interventions may not allow sufficient time for sustained glycemic changes to manifest in HbA1c values, whereas FBG is more sensitive to acute and short-term changes in glucose metabolism. These observations are, however, exploratory given that the meta-regression model for HbA1c was saturated, and should be examined in future reviews with a larger body of evidence. Previous studies have shown that cultural beliefs and practices significantly impact diabetes self-management behaviors in West African communities (42–44). Additionally, limited healthcare resources and access to trained professionals pose challenges to effective diabetes management in the region (45).

The substantial variability observed in our analysis may also stem from differences in intervention intensity, implementation fidelity, participant engagement, and adherence. A systematic review by Norris et al (14) emphasized that the effectiveness of self-management education varies widely, partly due to differences in program delivery and participant adherence. Interventions with frequent, structured sessions and clear implementation protocols tend to produce better outcomes, whereas those with less intensive or irregular programs may be less effective. Moreover, disparities in socioeconomic status, educational attainment, and health literacy levels in West Africa may influence participants ability to adhere to lifestyle interventions (45–47), further contributing to variability in the effectiveness of interventions. The heterogeneity observed in this review is likely not solely statistical but also conceptual, reflecting differences in intervention types, delivery strategies, and study contexts. Rather than representing random variation, this variability underscores the diversity of approaches used in real-world settings across West Africa. Consequently, the findings should be interpreted as evidence of a range of possible effects rather than a single uniform intervention impact.

Our study provides one of the first region-specific quantitative syntheses of lifestyle interventions for diabetes management in West Africa, offering contextually relevant insights that are often obscured in broader LMIC analyses.

However, this review has several important limitations. First, the small number of included studies limited the statistical power of meta-analyses and precluded robust subgroup and meta-regression analyses. Second, substantial heterogeneity, particularly for HbA1c outcomes, reduces confidence in pooled estimates and suggests that intervention effects are highly context-dependent. Third, variation in intervention design, intensity, and implementation fidelity was not consistently reported, limiting our ability to assess key drivers of effectiveness. Fourth, inclusion of quasi-experimental designs introduces potential bias and limits causal inference. Finally, restricting inclusion to English and French publications may have excluded relevant studies and introduced publication bias.

## Conclusion

Lifestyle-targeted interventions, specifically supervised physical activity, dietary modifications, and community-based diabetes education, alone or in combination, appear to improve glycemic control among adults with type 2 diabetes in West Africa, with more consistent evidence observed for FBG outcomes. The pooled mean difference of −1.81 mmol/L for FBG and −1.044% for HbA1c indicate clinically meaningful improvements, though the exceptional heterogeneity observed for HbA1c outcomes limits the interpretability of the pooled estimate. The impact of these interventions was not uniform across studies, and differences in sample size, intervention type, and duration did not fully explain the observed variability, suggesting that contextual factors including cultural beliefs, dietary habits, healthcare access, participant adherence, and intervention intensity may play a role, though these could not be directly assessed due to data limitations. Future research should prioritize larger, well-designed trials with standardized intervention protocols, improved reporting of adherence and implementation fidelity, and explicit attention to how local healthcare structures and social contexts can be leveraged to maximise the impact of lifestyle interventions in West Africa.

## Supporting information

S1 File. PRISMA Checklist

S1 Table. Characteristics and Quality Assessment of Studies

S2. File. Search Strategy

S3. File. Study Selection Process

## Data Availability

The minimal data set is available within the paper and its Supporting Information files

## Authors’ contributions

**Study protocol conceptualization:** Ellen Barnie Peprah (EBP)

**Study design**: Eugene Paa Kofi Bondzie (EPKB)

**Formal analysis**: Eugene Paa Kofi Bondzie (EPKB)

**Methodology:** Eugene Paa Kofi Bondzie (EPKB), Ellen Barnie Peprah (EBP), Yasmin Jahan (YJ)

**Supervision**: Dina Balabanova (DB), Irene Agyepong (IA), Tolib Mirzoev (TM)

**Validation**: Irene Agyepong (IA), Tolib Mirzoev (TM).

**Original draft writing**: Eugene Paa Kofi Bondzie (EPKB)

**Review and editing**: Eugene Paa Kofi Bondzie (EPKB), Nhyira Yaw Adjei-Banuah (NYB), Nana Efua Enyimayew Afun (EEA), Yasmin Jahan (YJ), Irene Agyepong (IA), Tolib Mirzoev (TM)

## Acknowledgments

We would like to acknowledge the London School of Hygiene and Tropical Medicine (LSHTM) public health library for support in retrieving some studies that were not easily accessible online.

## Supporting Information

**S1 File. PRISMA-P Checklist**

**S2 File. Study Selection Flowchart**

**S3 File. Study Selection Process**

**S1 Table. Characteristics and Quality Assessment of Studies**

## References

1. IDF_Atlas_10th_Edition_2021.pdf [Internet]. [cited 2023 Jun 19]. Available from: https://diabetesatlas.org/idfawp/resource-files/2021/07/IDF_Atlas_10th_Edition_2021.pdf

2. Diabetes [Internet]. [cited 2025 Feb 28]. Available from: https://www.who.int/news-room/fact-sheets/detail/diabetes

3. Harding JL, Pavkov ME, Magliano DJ, Shaw JE, Gregg EW. Global trends in diabetes complications: a review of current evidence. Diabetologia. 2019 Jan 1;62(1):3–16.

4. Ofori-Asenso R, Agyeman AA, Laar A, Boateng D. Overweight and obesity epidemic in Ghana—a systematic review and meta-analysis. BMC Public Health. 2016 Dec;16(1):1239.

5. Mensah GA, Roth GA, Fuster V. The Global Burden of Cardiovascular Diseases and Risk Factors: 2020 and Beyond. Journal of the American College of Cardiology. 2019 Nov 19;74(20):2529–32.

6. Olanrewaju YA, Oladunni AA, David KB, Babatunde YO, Damilola IA, Adedeji O, et al. Covid-19 and non-communicable diseases (NCDs) ian Africa: a narrative review. Afr Health Sci. 2023 Sep;23(3):412–21.

7. Basa M, Vries JD, McDonagh D, Comiskey C. The impact of COVID-19 on non-communicable disease patients in sub-Saharan African countries: A systematic review. PLOS ONE. 2024 Jun 21;19(6):e0293376.

8. The Global Diabetes Compact: a promising first year [Internet]. [cited 2023 Jun 19]. Available from: https://www.who.int/news/item/14-04-2022-the-global-diabetes-compact-a-promising-first-year

9. ElSayed NA, Aleppo G, Aroda VR, Bannuru RR, Brown FM, Bruemmer D, et al. 2. Classification and Diagnosis of Diabetes: Standards of Care in Diabetes—2023. Diabetes Care. 2023 Jan;46(Suppl 1):S19–40.

10. Colberg SR, Sigal RJ, Yardley JE, Riddell MC, Dunstan DW, Dempsey PC, et al. Physical Activity/Exercise and Diabetes: A Position Statement of the American Diabetes Association. Diabetes Care. 2016 Nov;39(11):2065–79.

11. Sigal RJ, Kenny GP, Wasserman DH, Castaneda-Sceppa C, White RD. Physical Activity/Exercise and Type 2 Diabetes: A consensus statement from the American Diabetes Association. Diabetes Care. 2006 Jun 1;29(6):1433–8.

12. Evert AB, Dennison M, Gardner CD, Garvey WT, Lau KHK, MacLeod J, et al. Nutrition Therapy for Adults With Diabetes or Prediabetes: A Consensus Report. Diabetes Care. 2019 May;42(5):731–54.

13. Umpierre D, Ribeiro PAB, Kramer CK, Leitão CB, Zucatti ATN, Azevedo MJ, et al. Physical Activity Advice Only or Structured Exercise Training and Association With HbA1c Levels in Type 2 Diabetes: A Systematic Review and Meta-analysis. JAMA. 2011 May 4;305(17):1790–9.

14. Norris SL, Nichols PJ, Caspersen CJ, Glasgow RE, Engelgau MM, Jack Jr. L, et al. Increasing diabetes self-management education in community settings: A systematic review. American Journal of Preventive Medicine. 2002;22(Suppl4):39–66.

15. Warburton DER, Nicol CW, Bredin SSD. Health benefits of physical activity: the evidence. CMAJ. 2006 Mar 14;174(6):801–9.

16. Greaves CJ, Sheppard KE, Abraham C, Hardeman W, Roden M, Evans PH, et al. Systematic review of reviews of intervention components associated with increased effectiveness in dietary and physical activity interventions. BMC Public Health. 2011 Feb 18;11(1):119.

17. Sagastume D, Siero I, Mertens E, Cottam J, Colizzi C, Peñalvo JL. The effectiveness of lifestyle interventions on type 2 diabetes and gestational diabetes incidence and cardiometabolic outcomes: A systematic review and meta-analysis of evidence from low- and middle-income countries. eClinicalMedicine [Internet]. 2022 Nov 1 [cited 2023 Jun 19];53. Available from: https://www.thelancet.com/journals/eclinm/article/PIIS2589-5370(22)00380-7/fulltext

18. Munan M, Dyck RA, Houlder S, Yardley JE, Prado CM, Snydmiller G, et al. Does Exercise Timing Affect 24-Hour Glucose Concentrations in Adults With Type 2 Diabetes? A Follow Up to the Exercise-Physical Activity and Diabetes Glucose Monitoring Study. Canadian Journal of Diabetes. 2020 Dec 1;44(8):711–718.e1.

19. Peprah E, Abdul-Basit AS, Appiah TD, Mirzoev T, Antwi E, Jahan Y, et al. Effectiveness of lifestyle interventions for glycemic control among adults with Type 2 Diabetes in West Africa: a systematic review and meta-analysis protocol. 2024 [cited 2024 Jan 22]; Available from: https://www.researchsquare.com/article/rs-3502674/latest

20. Page MJ, McKenzie JE, Bossuyt PM, Boutron I, Hoffmann TC, Mulrow CD, et al. The PRISMA 2020 statement: an updated guideline for reporting systematic reviews. Systematic Reviews. 2021 Mar 29;10(1):89.

21. Barker TH, Stone JC, Sears K, Klugar M, Leonardi-Bee J, Tufanaru C, et al. Revising the JBI quantitative critical appraisal tools to improve their applicability: an overview of methods and the development process. JBI Evidence Synthesis. 2023 Mar;21(3):478.

22. Braga F, Dolci A, Mosca A, Panteghini M. Biological variability of glycated hemoglobin. Clinica Chimica Acta. 2010 Nov 11;411(21):1606–10.

23. American Diabetes Association. 6. Glycemic Targets: Standards of Medical Care in Diabetes—2021. Diabetes Care. 2020 Dec 4;44(Supplement_1):S73–84.

24. Lenters-Westra E, Schindhelm RK, Bilo HJG, Groenier KH, Slingerland RJ. Differences in interpretation of haemoglobin A1c values among diabetes care professionals. Neth J Med. 2014 Nov;72(9):462–6.

25. Eyth E, Naik R. Hemoglobin A1C. In: StatPearls [Internet]. Treasure Island (FL): StatPearls Publishing; 2025 [cited 2025 Feb 26]. Available from: http://www.ncbi.nlm.nih.gov/books/NBK549816/

26. Higgins JPT, Altman DG, Gøtzsche PC, Jüni P, Moher D, Oxman AD, et al. The Cochrane Collaboration’s tool for assessing risk of bias in randomised trials. BMJ. 2011 Oct 18;343:d5928.

27. Borenstein M, Hedges LV, Higgins JPT, Rothstein HR. Introduction to meta-analysis [Internet]. wiley; 2009 [cited 2025 Feb 27]. Available from: http://www.scopus.com/inward/record.url?scp=84889351499&partnerID=8YFLogxK Schünemann HJ, Higgins JPT, Vist GE, Glasziou P, Akl EA, Skoetz N, Guyatt GH. Chapter 14: Completing ‘Summary of findings’ tables and grading the certainty of the evidence [last updated August 2023]. In: Higgins JPT, Thomas J, Chandler J, Cumpston M, Li T, Page MJ, Welch VA (editors). Cochrane

28. Handbook for Systematic Reviews of Interventions version 6.5. Cochrane, 2024. Available from www.training.cochrane.org/handbook.

29. Osho O, SRA A, Osinubi A, Olawale O. Effect of Progressive Aerobic and Resistance Exercises on the Pulmonary functions of Individuals with Type 2 Diabetes in Nigeria. Int J Endocrinol Metab. 2012 Mar 1;10:411–7.

30. Asuako B, Moses MO, Eghan BA, Sarpong PA. Fasting plasma glucose and lipid profiles of diabetic patients improve with aerobic exercise training. Ghana Med J. 2017 Sep;51(3):120–7.

31. Amin M, Kerr D, Atiase Y, Samir MM, Driscoll A. Improving Metabolic Syndrome in Ghanaian Adults with Type 2 Diabetes through a Home-Based Physical Activity Program: A Feasibility Randomised Controlled Trial. Int J Environ Res Public Health. 2023 Apr 14;20(8):5518.

32. Ezema CI, Omeh E, Onyeso OKK, Anyachukwu CC, Nwankwo MJ, Amaeze A, et al. The Effect of an Aerobic Exercise Programme on Blood Glucose Level, Cardiovascular Parameters, Peripheral Oxygen Saturation, and Body Mass Index among Southern Nigerians with Type 2 Diabetes Mellitus, Undergoing Concurrent Sulfonylurea and Metformin Treatment. Malays J Med Sci. 2019 Sep;26(5):88–97.

33. Fayehun AF, Olowookere OO, Ogunbode AM, Adetunji AA, Esan A. Walking prescription of 10 000 steps per day in patients with type 2 diabetes mellitus: a randomised trial in Nigerian general practice. Br J Gen Pract. 2018 Feb;68(667):e139–45.

34. C Afiaenyi I, K Ngwu E, M Okafor A, Ayogu RN. Effects of Moringa oleifera leaves on the blood glucose, blood pressure, and lipid profile of type 2 diabetic subjects: A parallel group randomized clinical trial of efficacy. Nutr Health. 2023 May 25;2601060231176873.

35. Essien O, Otu A, Umoh V, Enang O, Hicks JP, Walley J. Intensive Patient Education Improves Glycaemic Control in Diabetes Compared to Conventional Education: A Randomised Controlled Trial in a Nigerian Tertiary Care Hospital. PLoS One. 2017 Jan 3;12(1):e0168835.

36. Debussche X, Besançon S, Balcou-Debussche M, Ferdynus C, Delisle H, Huiart L, et al. Structured peer-led diabetes self-management and support in a low-income country: The ST2EP randomised controlled trial in Mali. PLoS One. 2018 Jan 22;13(1):e0191262.

37. R.T I, Kolawole B, E.O O, Salawu A, Ajose O, S A, et al. A Controlled Comparison of the Effect of a High Fiber Diet on the Glycaemic and Lipid Profile of Nigerian Clinic Patients with Type 2 Diabetes. Pakistan Journal of Nutrition. 2007 Feb 1;6.

38. Adeniyi AF, Uloko AE, Ogwumike OO, Sanya AO, Fasanmade AA. Time course of improvement of metabolic parameters after a 12 week physical exercise programme in patients with type 2 diabetes: the influence of gender in a Nigerian population. Biomed Res Int. 2013;2013:310574.

39. O’Donoghue G, O’Sullivan C, Corridan I, Daly J, Finn R, Melvin K, et al. Lifestyle Interventions to Improve Glycemic Control in Adults with Type 2 Diabetes Living in Low-and-Middle Income Countries: A Systematic Review and Meta-Analysis of Randomized Controlled Trials (RCTs). Int J Environ Res Public Health. 2021 Jun 10;18(12):6273.

40. Esposito K, Maiorino MI, Bellastella G, Chiodini P, Panagiotakos D, Giugliano D. A journey into a Mediterranean diet and type 2 diabetes: a systematic review with meta-analyses. BMJ Open. 2015 Aug 1;5(8):e008222.

41. Attridge M, Creamer J, Ramsden M, Cannings-John R, Hawthorne K. Culturally appropriate health education for people in ethnic minority groups with type 2 diabetes mellitus - Attridge, M – 2014 | Cochrane Library. [cited 2025 Feb 27]; Available from: https://www.cochranelibrary.com/cdsr/doi/10.1002/14651858.CD006424.pub3/information

42. Kangmennaang J, Siiba A, Dassah E, Kansanga M. The role of social support and the built environment on diabetes management among structurally exposed populations in three regions in Ghana. BMC Public Health. 2023 Dec 13;23(1):2495.

43. Bossman IF, Dare S, Oduro BA, Baffour PK, Hinneh TK, Nally JE. Patients’ knowledge of diabetes foot complications and self-management practices in Ghana: A phenomenological study. PLoS ONE. 2021 Aug 25;16(8):e0256417. doi: 10.1371/journal.pone.0256417.

44. Molla IB, Hagger V, Rothmann MJ, Rasmussen B. The Role of Community Organisation, Religion, Spirituality and Cultural Beliefs on Diabetes Social Support and Self-Management in Sub-Saharan Africa: Integrative Literature Review. J Relig Health [Internet]. 2025 Jan 24 [cited 2025 Feb 27]; Available from: 10.1007/s10943-024-02233-y

45. Suglo JN, Evans C. Factors influencing self-management in relation to type 2 diabetes in Africa: A qualitative systematic review. PLOS ONE. 2020 Oct 22;15(10):e0240938.

46. Mogre V, Abanga ZO, Tzelepis F, Johnson NA, Paul C. Adherence to and factors associated with self-care behaviours in type 2 diabetes patients in Ghana. BMC Endocrine Disorders. 2017 Mar 24;17(1):20.

47. Opoku R, Ackon SK, Kumah E, Botchwey COA, Appiah NE, Korsah S, et al. Self-care behaviors and associated factors among individuals with type 2 diabetes in Ghana: a systematic review. BMC Endocrine Disorders. 2023 Nov 22;23(1):256.

