## Supplementary material for "Effectiveness of Lifestyle Interventions for Glycemic Control among Adults with Type 2 Diabetes in West Africa: a Systematic Review and Meta-analysis": S2. File. Search Strategy

### SEARCH CONCEPT

#### Concept 1( Intervention)

"Diet Therapy"[Mesh] OR "Diet" OR "nutrition" OR “nutrition education” OR “nutrition counselling”

#### Concept 2 ( Intervention)

"Exercise"[Mesh] OR "Physical activit*”

#### Concept 3 (Context/Outcome)

"Diabetes Mellitus, Type 2"[Mesh] OR “Glycemic Control”[Mesh] OR diabetes OR "high blood glucose" OR "high blood sugar"

#### Concept 4 ( Context)

africa, western[MeSH Terms] West Africa OR “Benin” OR “Burkina Faso” OR “Cape Verde” OR Gambia OR Ghana OR Guinea OR Guinea-Bissau OR “Ivory Coast” OR Liberia OR Mali OR Mauritania OR Niger OR Nigeria OR Senegal OR Sierra Leone OR Togo

**Table 1 : PUBMED SEARCH STRATEGY**

| | |
|---|---|
| **#1** | "Diet Therapy"[Mesh] OR "Diet" OR "nutrition" OR “nutrition education” OR “nutrition counselling” |
| **#2** | "Exercise"[Mesh] OR "Physical activit*” |
| **#3** | "lifestyle behavio*" OR "lifestyle modification" |
| **#4** | "Diabetes Mellitus, Type 2"[Mesh] OR “Glycemic Control”[Mesh] OR diabetes OR "high blood glucose" OR "high blood sugar" |
| **( #1 OR #2 OR #3 ) AND #5** | |

**Sample search input for Pubmed**

("Diet Therapy"[MeSH Terms] OR "Diet"[All Fields] OR "nutrition"[All Fields] OR "nutrition education"[All Fields] OR "nutrition counselling"[All Fields]) AND ("Diet Therapy"[MeSH Terms] OR "Diet"[All Fields] OR "nutrition"[All Fields] OR "nutrition education"[All Fields] OR "nutrition counselling"[All Fields] OR ("Exercise"[MeSH Terms] OR "physical activit*"[All Fields]) OR ("lifestyle behavio*"[All Fields] OR "lifestyle modification"[All Fields])) AND ("diabetes mellitus, type 2"[MeSH Terms] OR "Glycemic Control"[MeSH Terms] OR ("diabete"[All Fields] OR "diabetes mellitus"[MeSH Terms] OR ("diabetes"[All Fields] AND "mellitus"[All Fields]) OR "diabetes mellitus"[All Fields] OR "diabetes"[All Fields] OR "diabetes insipidus"[MeSH Terms] OR ("diabetes"[All Fields] AND "insipidus"[All Fields]) OR "diabetes insipidus"[All Fields] OR "diabetic"[All Fields] OR "diabetics"[All Fields] OR "diabets"[All Fields]) OR "high blood glucose"[All Fields] OR "high blood sugar"[All Fields]) AND (("africa, western"[MeSH Terms] AND ("africa, western"[MeSH Terms] OR ("africa"[All Fields] AND "western"[All Fields]) OR "western africa"[All Fields] OR ("west"[All Fields] AND "africa"[All Fields]) OR "west africa"[All Fields])) OR "Benin"[All Fields] OR "Burkina Faso"[All Fields] OR "Cape Verde"[All Fields] OR ("gambia"[MeSH Terms] OR "gambia"[All Fields] OR "gambia s"[All Fields]) OR ("ghana"[MeSH Terms] OR "ghana"[All Fields] OR "ghana s"[All Fields]) OR ("guinea"[MeSH Terms] OR "guinea"[All Fields] OR "guinea s"[All Fields] OR "guineas"[All Fields]) OR ("guinea bissau"[MeSH Terms] OR "guinea bissau"[All Fields] OR ("guinea"[All Fields] AND "bissau"[All Fields]) OR "guinea bissau"[All Fields]) OR "Ivory Coast"[All Fields] OR ("liberia"[MeSH Terms] OR "liberia"[All Fields] OR "liberia s"[All Fields]) OR ("mali"[MeSH Terms] OR "mali"[All Fields]) OR ("mauritania"[MeSH Terms] OR "mauritania"[All Fields]) OR ("niger"[MeSH Terms] OR "niger"[All Fields]) OR ("nigeria"[MeSH Terms] OR "nigeria"[All Fields] OR "nigeria s"[All Fields]) OR ("senegal"[MeSH Terms] OR "senegal"[All Fields] OR "senegal s"[All Fields]) OR "Sierra Leone"[All Fields] OR ("togo"[MeSH Terms] OR "togo"[All Fields]))
