## Supplementary figures and images for "Effectiveness of Lifestyle Interventions for Glycemic Control among Adults with Type 2 Diabetes in West Africa: a Systematic Review and Meta-analysis"

### S3. File. Study Selection Process

Figure 2: Decision-making flowchart for screening

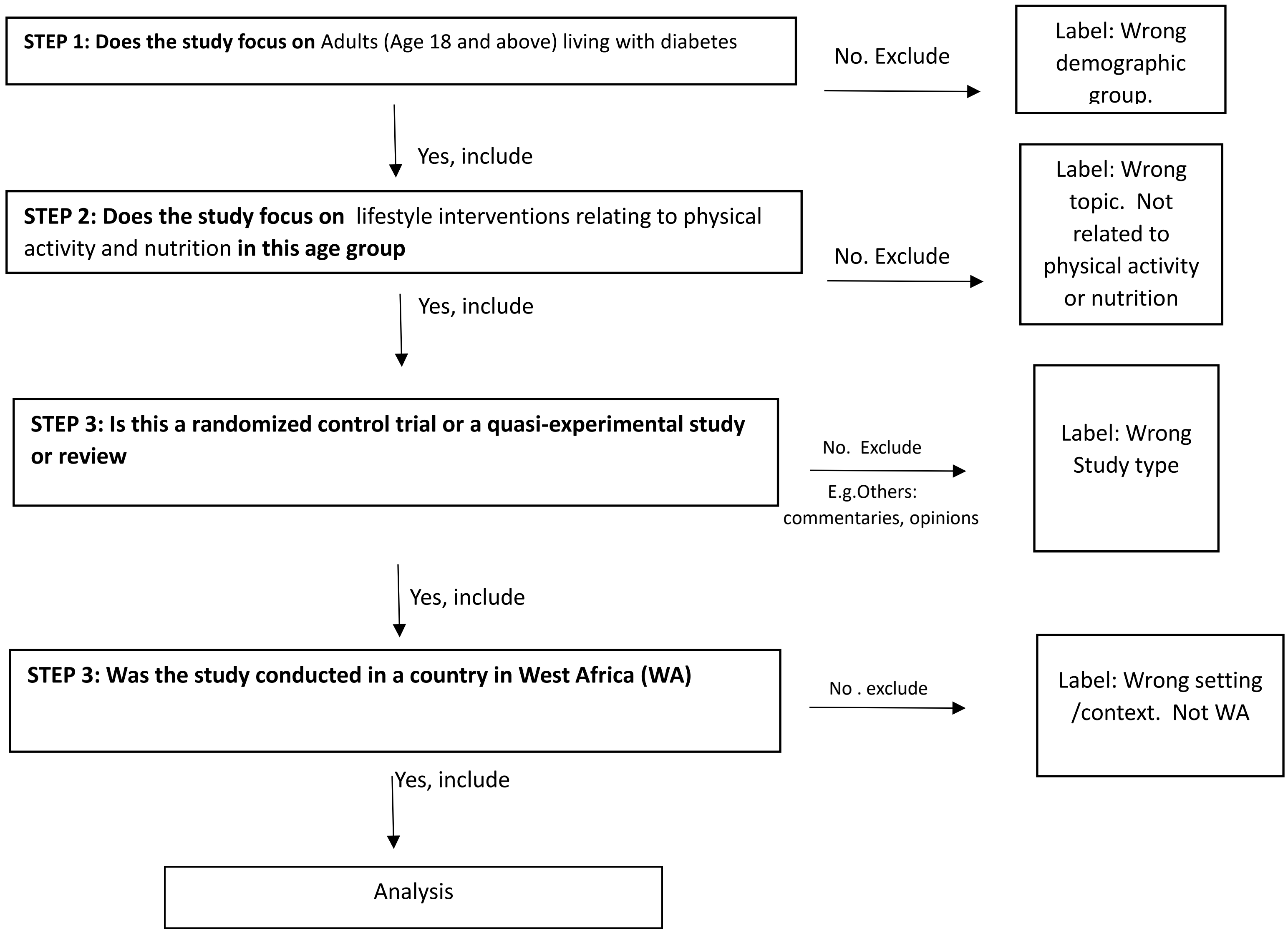
